# Modelling Decay of Population Immunity With Proposed Second Dose Deferral Strategy

**DOI:** 10.1101/2021.01.05.21249293

**Authors:** Jurgens Graham

## Abstract

A second dose deferred strategy has been proposed to increase initial population immunity as an alternative to the default two dose vaccine regimen with spacing of 21 or 28 days between vaccine doses for the mRNA vaccines from Pfizer and Moderna. This increased initial population immunity is only of value if one dose immunity does not decay so fast as to nullify the benefit. Because decay rates of one dose and two dose efficacy are currently unknown, a model to project population immunity between the two strategies was created. By evaluating the decay rate of one dose efficacy, two dose efficacy, and time until the second dose is given, the model shows that if there is an increased decay rate of one dose efficacy relative to the two dose decay rate, it is highly unlikely to nullify the benefit of increased population immunity seen in a second dose deferral strategy. Rather, all reasonable scenarios strongly favour a second dose deferral strategy with much higher projected population immunity in comparison to the default regimen.

## Modelling Decay of Population Immunity With Proposed Second Dose Deferral Strategy

### Rationale

The new mRNA vaccines from both Pfizer/BioNTech and Moderna have data showing a one dose efficacy rate of around 92% when data is collected starting 14 days after inoculation. For a population of any given risk bracket, deferral of the second dose of the new mRNA vaccine allows for an increased population immunity in a time of vaccine shortage. Such population immunity is key to preventing unnecessary loss of life and hospitalizations, now more important than ever given the increased surge of COVID-19 infections nationally. Hesitation to deviate from the manufacturer’s trial protocol appears to be primarily driven by concern about waning immunity to one dose of the vaccine if the second dose is delayed. Presented here is a modelling attempt to both quantify and visualize how an increased decay rate for such a second dose deferral strategy might affect population immunity.

**Image 1:**
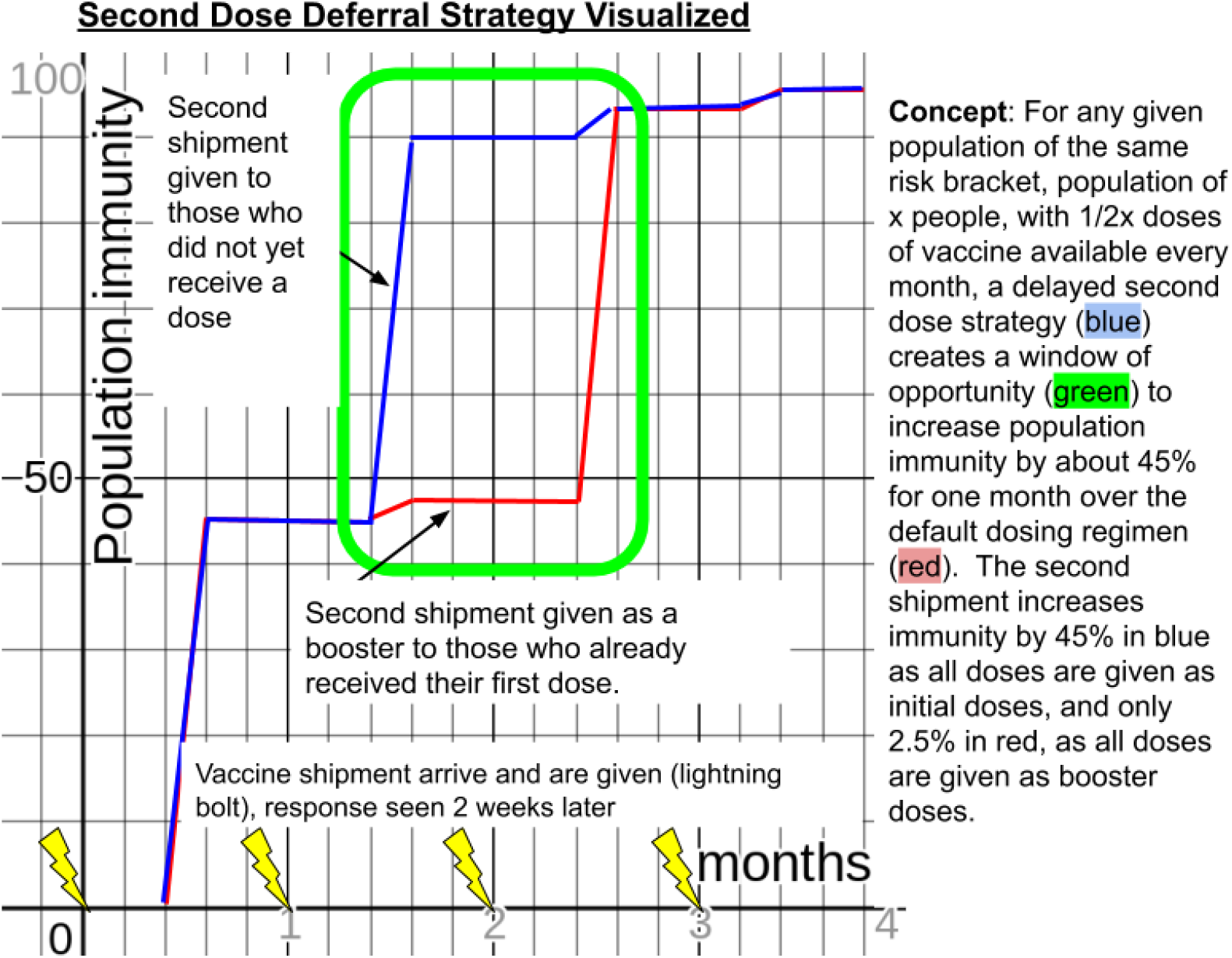
Concept of second dose deferred strategy: Second dose deferred strategy clearly increases initial and average population immunity. Any time an individual receives a second dose of a vaccine before everyone else in their risk bracket has received their first dose, the vaccine is not being distributed to optimize population immunity. The possible risk is that the decay rate after one dose may nullify the effects of that initial increase. (This graph assumes a population of the same risk bracket, ie/ all long term care residents, or all frontline health care workers, etc. Waning decay of immunity for one or two dose regimens are not visualized here, as they will be evaluated in further images. The variation of time until the second dose is available will also be further evaluated).

### Process

Data for duration of immunity with the mRNA vaccines is quite scarce at this point. The two dose regimen spaced apart 3 or 4 weeks appears to have 3 or 4 months of duration, and monitoring is ongoing. Two dose efficacy is 95% with a fairly high level of precision. One dose efficacy appears to be 92% starting 14 days after immunization with a wide 95% confidence interval of ∼69 to ∼98%. Single dose efficacy has been monitored for one week with Pfizer’s data and 2 weeks with Moderna’s data.

This model is predicated on the fact that we are in a vaccine shortage, with only one dose to give to each individual for a population. As illustrated in the above graph, the area the model is going to focus on is the green window of opportunity where in the default regimen individuals would have received their booster, while in the second dose deferred model those vaccine doses would be given to people in the same risk bracket who had not yet received a vaccination. Thus population immunity will start at 47.5% for the default regimen, Group A (only half the population is immunized with 2 doses for a 95% efficacy while the other half will receive no vaccination, 95%x0.5 +0%x0.5= 47.5%), and 90% for the second dose deferred strategy, Group B (everyone has received one dose).

Once individuals in a given risk bracket start receiving a second dose of vaccine without having every other individual in their risk bracket getting their first dose, then the vaccine is being used suboptimally from an initial population immunity perspective. If the average benefit from a booster is only 5%, (increasing efficacy from 90 to 95%), then intuitively it seems logical that this dose is better used by giving it to another individual without any vaccination to increase their immunity from 0 to 90%. However, to ensure intuition is in agreement with reality, the evaluation of decay rates of immunity is important to assess if potential increased decay with one dose could nullify the benefit in overall population immunity. The goal of the model is to help optimize our distribution strategy based on evidence rather than on intuition.

This model will evaluate 3 important variables that can diminish or nullify the expected benefit in population immunity obtained from a second dose deferred strategy: rate of decay of the vaccine after the default 2 dose regimen, rate of decay after a single dose (with increased spacing to a deferred second dose) relative to the decay rate after 2 doses, and the time until the second dose is given in the second dose deferred strategy.

#### Rate of decay of the vaccine after the default 2 dose regimen

The new mRNA vaccines have unknown duration of immunity and unknown kinetics of decay. A simple decay rate formula, y=ab^x is used here to estimate various levels of immunity with various decay rates. This formula is commonly used in SIR (susceptible, infected, resistant) modelling. To estimate the possible duration of immunity of SARS-Cov-2, a search was made to see the duration of immune response to infection from another coronavirus, in this case SARS-Cov-1. The immune response appears to be quite robust to SARS-Cov-1, lasting at least 11 years for T cell response (1). The SARS-Cov-1 immune response showed Th1 predominant CD4 and CD8 cells playing an important, if not primary, role in protection from reinfection, which is similar to the immune response elicited by the new mRNA vaccines. Using the SARS-Cov-1 immune response duration as a baseline for possible immunity, the decay rate was estimated as 0.5 to 1.5% per month for SARS immunity. Trying to err on the side of being conservative with the model, the faster decay rate of 1.5% was used, which would mean there would be 40% population immunity remaining to SARS at 5 years from immune response. GIven the robust response to the new mRNA vaccines with serological markers well above those who recovered from wild type infection of COVID-19, it is possible that this decay rate might lead to an underestimation of duration of immunity. Recovery after COVID-19 confers protection against reinfection for at least six months (2) and induces long-lived bone marrow cells (3). However, given the unknown decay rate, using the 1.5% decay rate per month seemed like a reasonable low end estimate.

There is some concern that the duration of immunity might be much shorter lived because common cold coronaviruses can cause reinfection even one year later, although most of the time the infections are asymptomatic or milder than initial infection (4). This indicates the possibility of a higher decay rate, in the range of 5% per month. Thus the model will evaluate an upper high end estimate of decay rate of 10%/month for the two dose vaccine. (From the cumulative incidence graphs on both Pfizer’s and Moderna’s phase 3 trial data, there does not appear to be much, if any, waning efficacy of the vaccine after 4 months, so the 10% estimate is likely much higher than realistic based on our current information.)

#### Rate of decay after a single dose relative to 2 doses

If there is an increased immunity of decay after receiving 2 doses, it is unknown how much longer lasting it might be compared to one dose. It is possible that the one dose immunity decay rate would be the same as two dose immunity decay. The body is responding to the same antigen, and if there is enough stimulation to have an efficacy of 90% after one dose, then the expansion of CD4 and CD8 cells and the antibodies created may decay at a similar rate as the 2 dose regimen, just starting at a level that is 5 or 10% less. However, considering the higher levels of immune stimulation after the second dose with CD4 and CD8 cells and higher antibody levels, a plausible scenario is that one dose decay rate might decay slightly quicker than the two dose regimen. The second dose may not only boost the overall efficacy of the vaccine by around 5-10%, but may also boost the duration of immunity by 5-10%. An increased decay rate of 0-25% seems most likely as an estimate for one dose immunity decay rate compared to two dose decay rate given the restimulation of the immune system with the second dose. However, to allow for the uncertainty in the model given current lack of data, one dose decay rates much higher were evaluated, such as 100% and even 500% increase relative to the decay rate of two doses.

#### Time until the second dose is given

The time until the second dose is given can vary depending on the size of the risk bracket the second dose deferred strategy is being applied to, and vaccine production. For high risk groups, being a smaller percentage of the population, the time needed for everyone to get their initial dose before having enough supply to start given second doses would likely not be more than 2 or 3 months. For low risk groups, a relatively larger percentage of the population, the time needed for everyone to get their initial dose before having enough supply to start with the second dose may be closer to 6 or 9 months, but generally not expected to be more than 12 months. The model will focus on time frames from 2 to 24 months.

### Modelling Results

Using a variety of decay rates and time frames, it is possible to see where the proposed strategy of second dose deferred regimen is preferred to the default 2 dose regimen and vice versa.

**Image 2:**
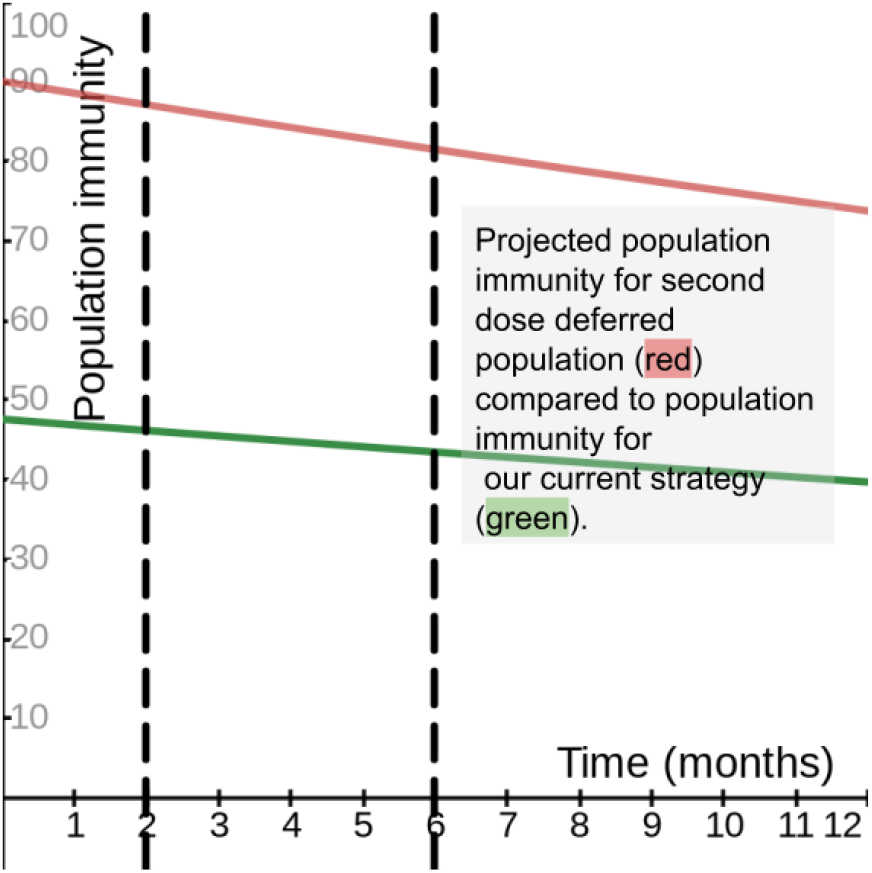
Scenario using reasonable estimates of each variable: Projected population immunity based on reasonable estimates of values for the variables. Decay rate of the default regimen is 1.5% per month and the decay rate of one dose efficacy wanes by 10% faster than two dose efficacy. Black vertical dashed lines show estimated time frames when a second dose would be available after everyone in a risk bracket has received their first vaccination.

**Image 3:**
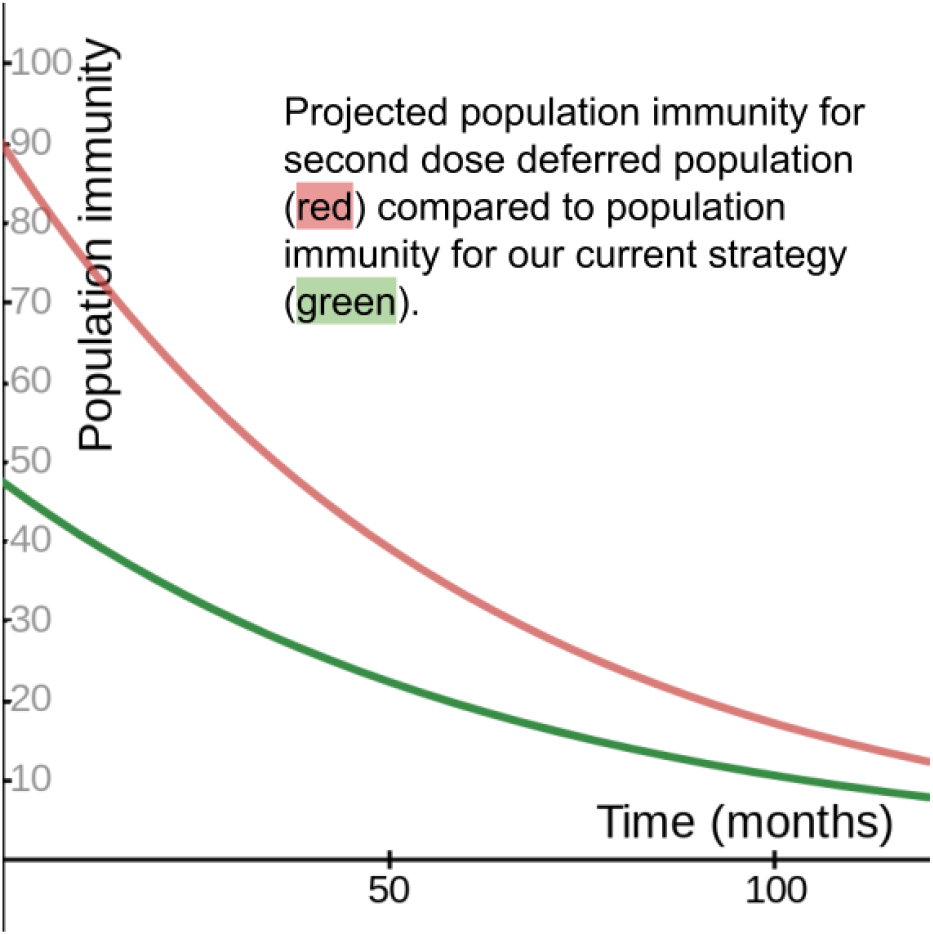
Zoomed out look at image 2: Evaluating an extended time frame with the same variables as above. Projected population immunity based on reasonable estimates of the variables, decay rate of the default regimen is 1.5% per month and the decay rate of one dose efficacy wanes by 10% faster. Even if it takes 10 years to have the second dose available, the second dose deferred strategy would clearly increase population immunity relative to the default strategy.

**Image 4:**
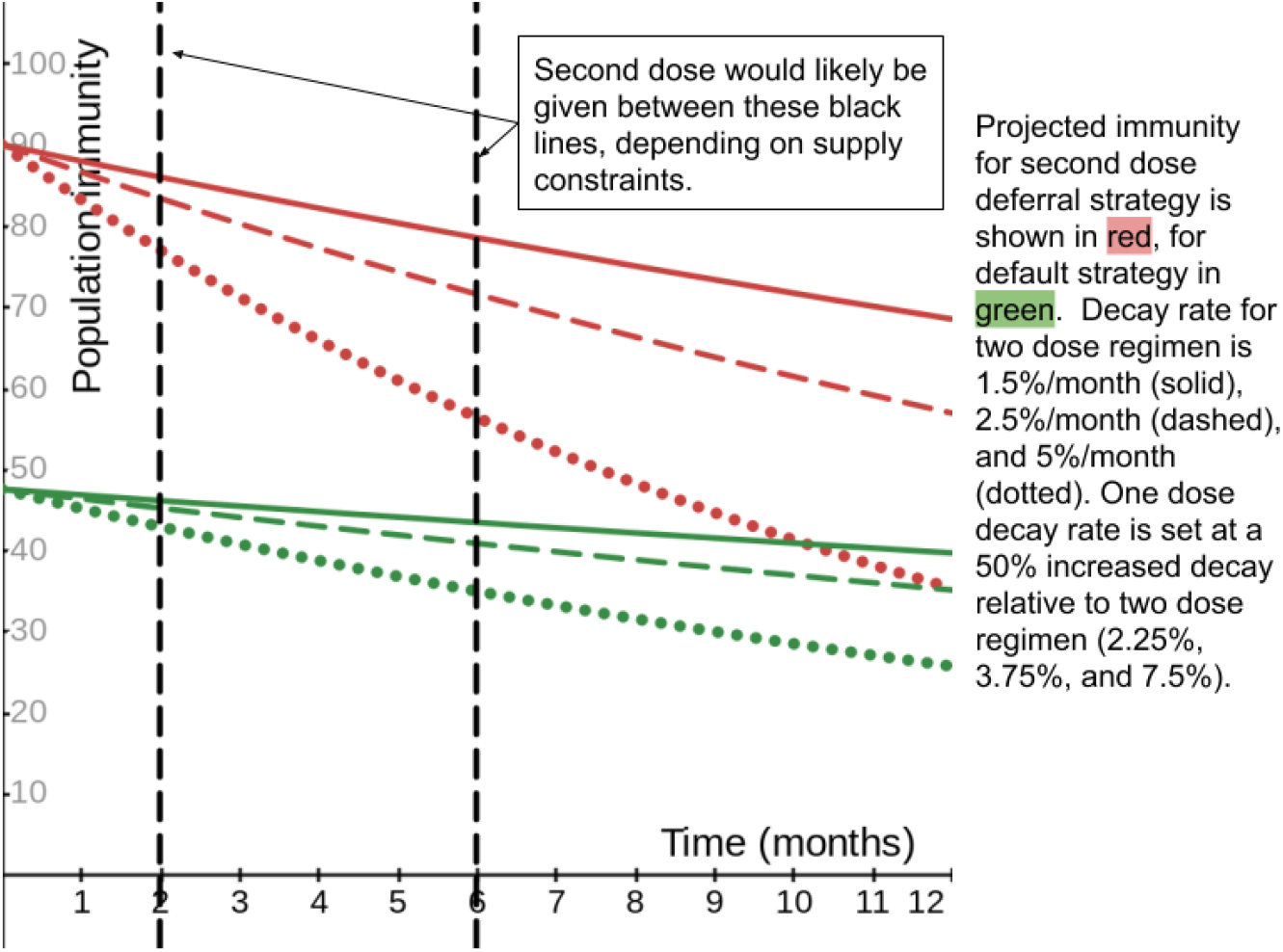
Evaluating varying decay rates of default strategy: The second dose deferral strategy (red) shows clear benefit to population immunity relative to the default regimen (green) with the default regimen decay rates varying from 1.5%/month to 5%/month and the relative increase in decay rate for one dose efficacy being set at 50% higher. The same type of line (solid, dashed or dotted) should be compared between the two strategies. Realistic time frames until a second dose might be available are shown as bordered by black dashed vertical lines.

**Image 5:**
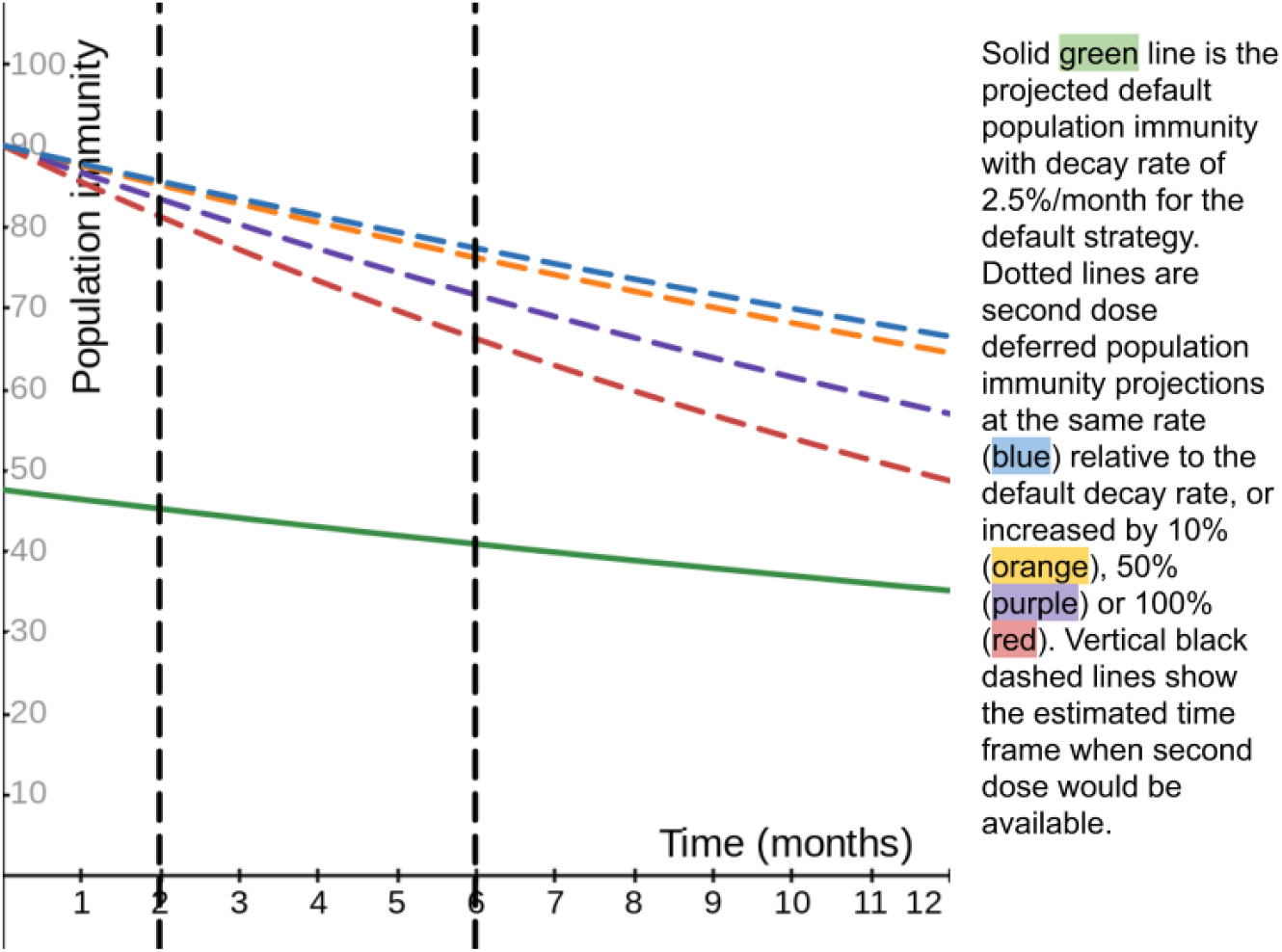
Evaluating varying decay rates of one dose efficacy relative to the default decay rate: The decay rate is set at 2.5% for the two dose regimen. The decay rate for one dose efficacy ranges from the same decay rate to 100% increased relative to the 2 dose regimen. Even with a decay rate increased 100% compared to the default regimen, the second dose deferral strategy clearly increases the population immunity.

**Image 6:**
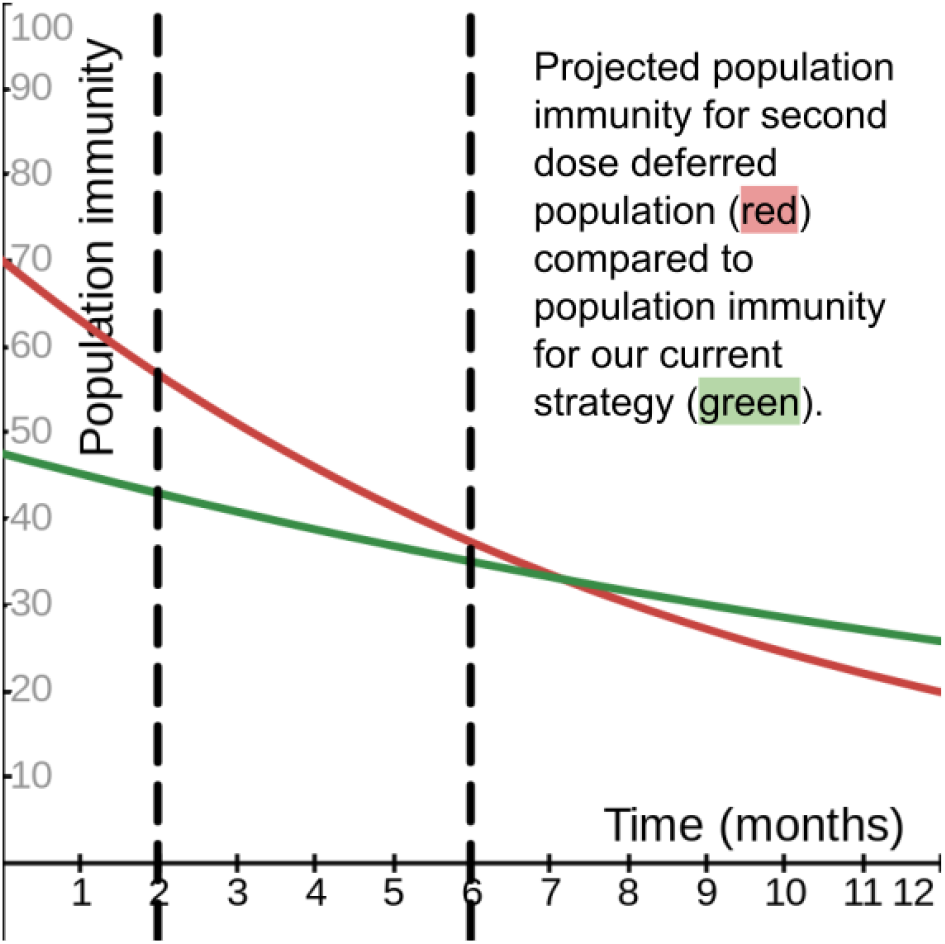
Evaluation using highly cautious estimation of variables: Projection of population immunities using decay rate of 5%/month for the default 2 dose regimen, shown in green, with a 100% relative increase in decay for one dose efficacy, shown in red. The population immunity for the second dose deferral strategy uses the lower limit of the 95% confidence interval of one dose efficacy at ∼70% (instead of using the most likely efficacy at 92% based on the data we have). Black vertical dashed lines show estimated time frames when a second dose would likely be available Note that after about 7 months the second dose deferral strategy population immunity is below the default strategy population immunity, but because the total area under the curve of red is greater than green, the second dose deferred strategy is still the preferred option. Even with such a conservative estimate of benefit for the second dose deferred strategy, the second dose deferred strategy is still clearly favoured over the default spacing regimen for more than 6 months.

**Table A to D:** Summary of findings: Population immunity decay rate estimates of two dose regimen compared with relative increase in one dose decay rates. Baseline decay rates for the two dose regimen are 1.5%, 2.5%, 5%, and 10% per month for A to D respectively. The one dose efficacy decay rate variable is shown in the left column (relative to the 2 dose decay rates) for each table. Areas highlighted in green are scenarios favored by the second dose deferred strategy. Yellow highlights favor the second dose deferred strategy with higher average population immunity (despite lower final population immunity). Figures highlighted in red favor the default strategy. Blue box indicates most reasonable estimates of time and relative decay rates. Red highlights scenarios that favor the current strategy. * Indicates equivalent average population immunity between the two strategies.

| Relative Increase of decay from two dose regimen to deferred second dose regimen | Default 2 dose regimen, decay rate at 1.5%/month, projected population immunity at 3, 6, 12, and 24 months | Second dose deferred strategy, decay rate as given in left column, projected population immunity at 3, 6, 12, and 24 months |
| --- | --- | --- |
| Equivalent decay rate (1.0) | 45, 43, 40, 33 | 86, 82, 75, 63 |
| Increased 10% (1.1 fold) | 45, 43, 40, 33 | 86, 81, 74, 60 |
| Increased 25% (1.25 fold) | 45, 43, 40, 33 | 85, 80, 72, 57 |
| Increased 50% (1.5 fold) | 45, 43, 40, 33 | 84, 79, 68, 52 |
| Increased 100% (2 fold) | 45, 43, 40, 33 | 82, 75, 62, 43 |
| Increased 5 fold | 45, 43, 40, 33 | 71, 56, 35, 14 |

C. Estimated 5% baseline decay rate of immunity for default regimen, 2 doses given 21 days apart
| Relative Increase of decay from two dose regimen to deferred second dose regimen | Default 2 dose regimen, decay rate at 5%/month, projected population immunity at 3, 6, 12, and 24 months | Second dose deferred strategy, decay rate as given in left column, projected population immunity at 3, 6, 12, and 24 months |
| --- | --- | --- |
| Equivalent decay rate (1.0) | 41, 35, 26, 14 | 77, 66, 49, 26 |
| Increased 10% (1.1 fold) | 41, 35, 26, 14 | 76, 64, 46, 23 |
| Increased 25% (1.25 fold) | 41, 35, 26, 14 | 74, 61, 41, 19 |
| Increased 50% (1.5 fold) | 41, 35, 26, 14 | 71, 56, 35, 14 |
| Increased 100% (2 fold) | 41, 35, 26, 14 | 66, 48, 25, 7 |
| 5 fold increase | 41, 35, 26, 14 | 38, 16*, 3, 0 |

B. Estimated 2.5% baseline decay rate of immunity for default regimen, 2 doses given 21 days apart
| Relative Increase of decay from two dose regimen to deferred second dose regimen | Default 2 dose regimen, decay rate at 2.5%/month, projected population immunity at 3, 6, 12, and 24 months | Second dose deferred strategy, decay rate as given in left column, projected population immunity at 3, 6, 12, and 24 months |
| --- | --- | --- |
| Equivalent decay rate (1.0) | 44, 41, 35, 26 | 83, 77, 66, 49 |
| Increased 10% (1.1 fold) | 44, 41, 35, 26 | 83, 76, 64, 46 |
| Increased 25% (1.25 fold) | 44, 41, 35, 26 | 82, 74, 61, 42 |
| Increased 50% (1.5 fold) | 44, 41, 35, 26 | 80, 72, 57, 36 |
| Increased 100% (2 fold) | 44, 41, 35, 26 | 77, 66, 49, 26 |
| Increased 5 fold | 44, 41, 35, 26 | 60, 40, 18*, 3 |

D. Estimated 10% baseline decay rate of immunity for default regimen, 2 doses given 21 days apart
| Relative Increase of decay from two dose regimen to deferred second dose regimen | Default 2 dose regimen, decay rate at 10%/month, projected population immunity at 3, 6, 12, and 24 months | Second dose deferred strategy, decay rate as given in left column, projected population immunity at 3, 6, 12, and 24 months |
| --- | --- | --- |
| Equivalent decay rate (1.0) | 35, 25, 13, 4 | 66, 48, 25, 7 |
| Increased 10% (1.1 fold) | 35, 25, 13, 4 | 63, 45, 22, 5 |
| Increased 25% (1.25 fold) | 35, 25, 13, 4 | 60, 40, 18, 4 |
| Increased 50% (1.5 fold) | 35, 25, 13, 4 | 55, 34, 13, 2 |
| Increased 100% (2 fold) | 35, 25, 13, 4 | 46, 24, 6, 0* |
| 5 fold increase | 35, 25, 13, 4 | 11, 1, 0, 0 |

## Discussion

In summary, modelling projected decay rates for a proposed second dose deferred strategy relative to the default 2 dose strategy decay rates can help guide vaccine distribution strategy. Based on the modelling as shown above, if future data shows there is an increased decay rate of one dose efficacy relative to two dose efficacy, it is highly unlikely it will nullify the increased population immunity of the deferred second dose strategy. In nearly every evaluated scenario the second dose deferred strategy was favored relative to the default 2 dose (spaced 21/28 days apart) strategy. Even using extreme estimates of the variables, the second dose deferral strategy often has higher average projected population immunity, and it is only in using a combination of extreme estimates with multiple variables that the default regimen would be favoured over the proposed second dose deferral regimen. From an immunology and physiological point of view there seems to be no apparent reason why the decay rate would be drastically higher with the one dose efficacy regimen relative to the default 2 dose regimen, especially considering the majority of the efficacy of the vaccine is obtained with the first dose. No current data available suggests such an extreme level of decay for one dose efficacy at this point. However, given that there still remains the possibility, minute though it may be, that the decay rate for one dose efficacy could indeed be severe, ongoing monitoring will be essential to evaluate ongoing efficacy of one dose efficiency, should the proposed deferred second dose strategy be implemented.

With what seems like clear benefit to delaying the second dose in population immunity until supply of vaccine is replenished, the next important question to address is if longer spacing between the two doses of the vaccine will decrease long term efficacy. Unfortunately, phase 1 and 2 trials did not address long term efficacy by testing spacing regimens at all. The 21 or 28 day spacing for the second dose was likely chosen to restimulate the immune system quickly rather than to optimize long term efficacy. Long term efficacy can certainly be affected by spacing between the doses, but most of the time it is spacing that is too close together that leads to suboptimal long term immunity and increased side effects, as the following quote from the CDC illustrates: “Doses administered too close together…can lead to a suboptimal immune response. Certain vaccines produce increased rates of local or systemic reactions in certain recipients when administered more frequently than recommended”. Looking at the minimum spacing for most of our other vaccines, a trend can be observed that minimum spacing is typically at least 4 weeks, with recommended spacing often 8 weeks or longer (5).

Though the vaccines are different, the immune system responsible for increased protection after stimulation remains the same, and given the lack of harm with further spacing with well-established regimens, it is most likely the immune system will respond in a similar manner to the new mRNA vaccines. Rather than decreased long term efficacy, it is plausible that increased spacing between the two doses of longer than the 21/28 days used in the protocol could actually increase the duration of efficacy of these new mRNA vaccines, possibly with lower side effects.

Based on the model shown, even delaying the second dose by 12 months is highly unlikely to nullify the benefit of increased population immunity. However, with implementation of a deferred second dose strategy, ongoing monitoring to monitor one dose efficacy decay would lead to much more precise estimates of immunity decay for one dose efficacy. That data will be able to guide with greater certainty what time frames can be used in deferring the second dose to optimize population immunity. Given the robust, yet imprecise, one dose efficacy of greater than 90%, it is possible that a second dose of the vaccine may not be beneficial at all, though a larger sample size of one dose efficacy is needed to give a more precise estimate of its efficacy. Given the higher immune markers noted after the second dose of vaccine in Phase 1 and 2 trials, it is reasonable to suspect the second dose will confer a 5-10% increase in both efficacy against infection as well as duration of immunity compared to a single dose.

## Conclusion

Increasing population immunity is key in preventing unnecessary loss of life and harm to the economy. Implementing a deferred second dose strategy appears to maximize the effect of the vaccine supply, leading to a significant increase in population immunity and the associated reduction in loss of life. By modelling various decay rates, we can be reassured that given the data available to us at this time, it is both rational and prudent to deviate from the manufacturers trial designs, which neither considered optimal distribution for the benefit of the population nor evaluated the effect of spacing on long term efficacy. Indeed, the benefit of the increased population immunity of the proposed deferred second dose regimen vastly outweighed the speculative risk of increased decay rates for one dose efficacy. Evaluation of immunity decay rates as shown in this model strongly support implementation of the proposed deferred second dose strategy, along with continued evaluation of one dose efficacy decay rates.

## Data Availability

Data self contained in submission

